# The microenvironment of ulcerated acral melanoma is characterised by an inflammatory milieu and an enhanced humoral immune response

**DOI:** 10.1101/2025.05.05.25325616

**Authors:** Martha Estefania Vázquez-Cruz, Patricia Basurto-Lozada, Christian Molina-Aguilar, Bram van Haastrecht, Irving Simonin-Wilmer, Sophia Orozco-Ruiz, Héctor Martinez-Said, Alethia Álvarez-Cano, Dorian Y. García-Ortega, Ingrid Ferreira, Alfredo Hidalgo-Miranda, Diego Hinojosa-Ugarte, Luis Alberto Tavares-de-la-Paz, Jonadab E Olguin, Isabella Salinas, Axel Rodríguez-Pérez, Julia M. Martínez-Gómez, Ivar T. van der Zee, David Robert Grimes, Julio Saez-Rodriguez, Helena Vidaurri de la Cruz, Julia A. Newton-Bishop, D. Timothy Bishop, Mitchell P. Levesque, Patrícia A. Possik, David J. Adams, Carla Daniela Robles-Espinoza

**Affiliations:** Laboratorio Internacional de Investigación sobre el Genoma Humano (LIIGH), UNAM, Mexico; Department of Computer Science, VU Bioinformatics, Vrije Universiteit Amsterdam, De Boelelaan 1111, 1081 HV, Amsterdam, The Netherlands; Licenciatura en Ciencias Genómicas, Escuela Nacional de Estudios Superiores Unidad Juriquilla, Universidad Nacional Autónoma de México, Querétaro, 76230, México; Surgical Oncology, Skin, Soft Tissue & Bone Tumours Department, National Cancer Institute, Mexico City, Mexico; Surgical Oncology, Christus Muguerza Alta Especialidad, Monterrey, Nuevo Leon, Mexico; Wellcome Sanger Institute, Hinxton, Cambridgeshire, CB10 1SA, UK; Laboratorio de Genómica del Cáncer, Instituto Nacional de Medicina Genómica (INMEGEN), Mexico City, Mexico; Surgical Oncology, Bajio Regional High Specialty Hospital, Leon, Mexico; Laboratorio Nacional en Salud Diagnóstico Molecular y Efecto Ambiental en Enfermedades Crónico-Degenerativas, Facultad de Estudios Superiores Iztacala, Universidad Nacional Autónoma de México, Av. de los Barrios No. 1, Los Reyes Iztacala, Tlalnepantla de Baz, Estado de México, Mexico; Krantz Family Center for Cancer Research, Department of Medicine, Massachusetts General Hospital; Broad Institute of MIT and Harvard; Department of Medicine, Harvard Medical School; Department of Dermatology, University Hospital Zurich, Gloriastrasse 31, 8091, Zurich, Switzerland; Division of Pharmacoepidemiology and Clinical Pharmacology, Utrecht Institute for Pharmaceutical Sciences Utrecht University, Utrecht The Netherlands; School of Physical Sciences, Dublin City University, Dublin, Ireland; Discipline of radiation therapy, Trinity College Dublin, Trinity Centre for Health Sciences, St. James’s Hospital, Dublin, Ireland; Institute for Computational Biomedicine, Faculty of Medicine, Heidelberg University, Germany; Hospital de Especialidades de la Ciudad de México Dr Belisario Domínguez, SEDESA, Mexico City, México; Leeds Institute of Medical Research, School of Medicine, University of Leeds, Leeds LS9 7TF, UK; Leeds Institute of Cancer and Pathology, University of Leeds, Leeds, UK; Division of Basic and Experimental Research, Brazilian National Cancer Institute, Rua Andre Cavalcanti 37, Rio de Janeiro, RJ, 20231-050, Brazil

**Keywords:** ulceration, acral, melanoma, immune, microenvironment, TME

## Abstract

Acral melanoma (AM) is a distinct and understudied subtype of melanoma that is reported to represent the majority of melanoma diagnoses in various Latin American, African and Asian countries. Patients with ulcerated AM diagnoses face a worse prognosis and increased risk of recurrence. While tumour ulceration has been shown to influence therapy response in cutaneous melanoma patients, the clinical and molecular traits of AM tumours remain poorly understood. In this study, we performed transcriptomic profiling of 59 AM samples and proteomic analysis of 45 AM samples from patients with extensively annotated clinical information. Our analysis revealed immunological differences in ulcerated tumours, including a significant upregulation of processes related to humoral immunity and markers for macrophages/monocytes, alongside a downregulation of keratins, epidermis-associated processes and cell adhesion. Notably, ulcerated tumours exhibited disruption of tight junctions and desmosomes, potentially explaining their compromised tissue integrity. We identified a significant increase of plasma cells, M0 macrophages and eosinophils within the ulcerated tumour microenvironment, suggesting that inflammation and infection might accompany these lesions. Moreover, fibronectin, CD8, PD-1, CD14 and CD68 protein levels were useful in distinguishing between ulcerated and non-ulcerated samples using a random forest classifier. These findings indicate that persistent inflammation and dysregulated immune responses characterise ulcerated AM, potentially offering new avenues for targeted therapeutic interventions in this aggressive melanoma subtype.

## Introduction

Acral melanoma (AM) is a subtype of melanoma that occurs in non-hair bearing skin such as the palms, soles, and nail bed^1^, and is associated with worse prognosis and higher risk of recurrence than other subtypes of melanoma^2–4^. In several countries in Latin America, Asia, and Africa, it is reported as the most frequently diagnosed subtype of melanoma, although it represents a small proportion of melanoma cases in individuals of European descent^5^. Due to this, it has been scarcely studied compared to other melanoma subtypes, and many questions remain regarding its causes, genomic drivers and prognostic factors^6–9^. A significant number of AM patients present with ulceration, defined as the loss of epidermal matrix including the stratum corneum and basement membrane^10^, having a worse disease-specific survival and an increased risk of recurrence and brain metastases^3,11,12^. Ulceration, therefore, is recognised as a negative prognostic factor in AM.

In non-acral cutaneous melanoma (CM), ulceration status can influence therapy response, with patients with ulcerated tumours responding better to adjuvant interferon (IFN) therapy^13,14^. Ulcerated CM also has distinct tumoural and clinical characteristics from non-ulcerated CM: (i) a higher degree of vascularisation^15^, (ii) distinct gene and protein expression patterns, including a higher expression of matrix metalloproteinases, loss of E-cadherin and *PTEN*, and (iii) higher expression of MHC class I molecules (reviewed in ^16^). Ulcerated CM has also been reported to have a higher infiltration of macrophages, dendritic cells and tumour cell PD-L1 expression^12^. This evidence points to an important role of the immune system in ulcerated tumours that can impact treatment response in CM. However, to the best of our knowledge, there are no studies focusing on elucidating gene and protein expression profile, as well as the patterns of immune infiltration, in ulcerated vs non-ulcerated AMs^17^. Here, we find that ulcerated AM tumours show an exacerbated inflammatory and humoral immune response through the study of the transcriptomes of 59 primary tumours, as well as proteomic data from 45 patients. In this work, we are focusing on the study of tumours from Mexican patients, a Latin American population underrepresented in genomic studies of cancer^18–20^.

## Results

### Description of patient cohort and clinical characteristics

Fifty-nine Mexican patients diagnosed with a pathologically confirmed AM were included in this study (**Supplementary Table 1**, **Methods**). We focus our analysis exclusively on primary tumours. Patients were classified into the ulcerated (n=36) and non-ulcerated (n=23) groups based on histopathological confirmation at the time of diagnosis. Most tumours were located on the feet (n=50, **Supplementary Table 1**). Patients in the ulcerated group were more frequently male (44% vs 17%, two-tailed Fisher exact test *P-*value=0.048), had more recurrences (64% vs 26%, two-tailed Fisher exact test *P-*value=0.0073) and tended to have more distant metastases (47% vs 28%, two-tailed Fisher exact test *P-*value=0.058). Patients with ulcerated tumours also showed a tendency to be older (Wilcoxon Mann Whitney [WMW] *P-*value=0.053, **Figure 1a**), and their tumours had higher Breslow depths (WMW *P-*value=1.2x10^-07^, **Figure 1b**) and mitotic indexes (*P-*value=8.6x10^-05^, **Figure 1c**) than non-ulcerated lesions. More advanced tumour stages also had increasingly higher proportions of ulcerated tumours (**Figure 1d**). These observations are in accordance with what has been reported in the literature for cutaneous melanoma^21^.

**Figure 1.**
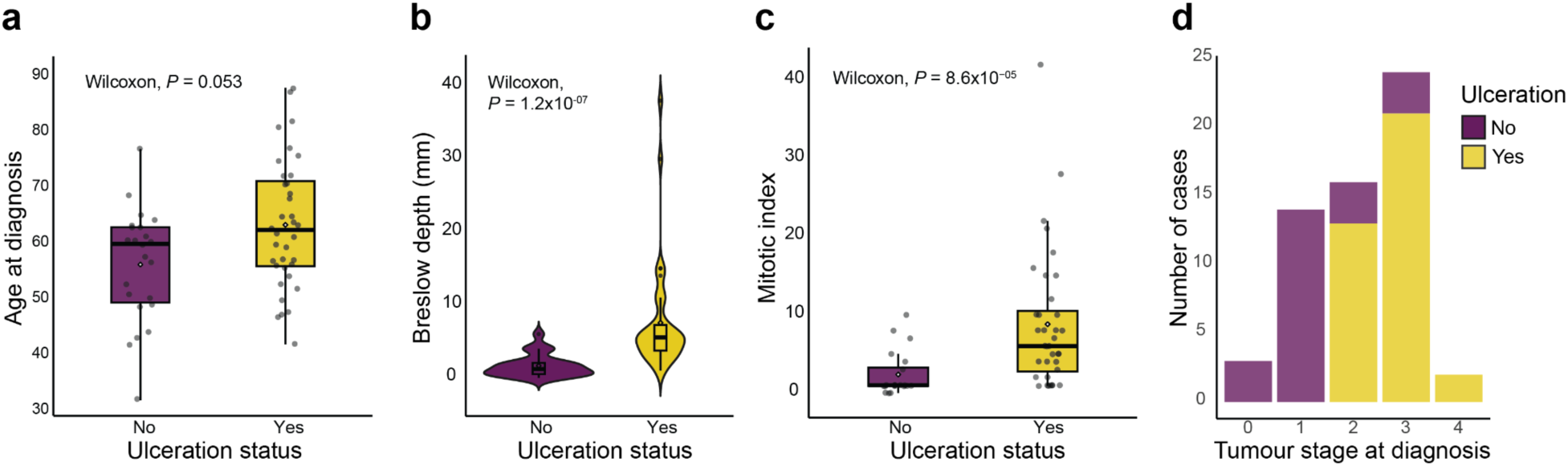
Clinical characteristics of AM patients according to ulceration status. Boxplots comparing differences in a) Age at diagnosis (P = 0.053), b) Breslow thickness (mm), and c) Mitotic index in ulcerated versus non-ulcerated AMs. d) Stacked bar charts showing the frequency of ulcerated tumours by stage. In all instances, yellow denotes ulcerated lesions, whilst purple represents non-ulcerated ones. Statistical significance was evaluated using the Wilcoxon rank-sum test.

### Differential gene expression and enrichment analyses show an upregulation of immunoglobulin-related processes and downregulation of cell-cell junctions in ulcerated tumours

We performed RNA extraction and sequencing from these 59 tumours (**Figure 2a, Supplementary Table 2**, **Methods**). To gain insights into the molecular differences between ulcerated and non-ulcerated tumours, we performed differential expression analysis controlling for sex, age and batch (**Figure 2b, Supplementary Table 3, Methods**). Gene set enrichment analysis (GSEA) revealed an increase in immune processes, including upregulation of genes like *POU2AF1*, *CD79A*, *MZB1*, and *FCRL5*, as well as immunoglobulin-related genes such as *IGHM*, *IGHG1*, *IGHG3*, *IGKC,* and other genes associated to adaptive immune-related responses. Conversely, there was a downregulation of keratins, epidermis-associated processes and cell adhesion in ulcerated tumours (**Figures 2c-d**, **Supplementary Table 4**). We observed an upregulation of vimentin and beta-catenin (**Supplementary Figure 1a**), as well as disruption of tight junctions and desmosomes in ulcerated samples, indicated by the downregulation of various epidermal and keratinocyte differentiation processes and intercellular junctions, including genes such as *CLDN1, CDH13, LAMB4,* protein junctional adhesion molecule-A *(JAM-A*) and desmosomal components such as *PERP*, *TP63*, *JUP* and *DSP* (**Supplementary Figures 1b,c**). To identify potential transcriptional regulators of these differentially expressed genes in ulcerated tumours, we performed transcription activity inference based on univariate linear model and prior knowledge networks (**Figure 2e, Supplementary Table 5**, **Methods**). Our results suggest that *ZEB1*, *E2F1, E2F4, FOXD1,* and *ATF6,* among others, are activated in the ulcerated group when compared to the non-ulcerated group, highlighting the proliferative and invasive phenotype active in these tumours. These results suggest that ulcerated samples exhibit an exacerbated humoral response, coupled to the activity of transcription factors -most notably *ZEB1*-that are associated with B cell differentiation and more aggressive tumoural characteristics (invasion and metastatic potential).

**Figure 2.**
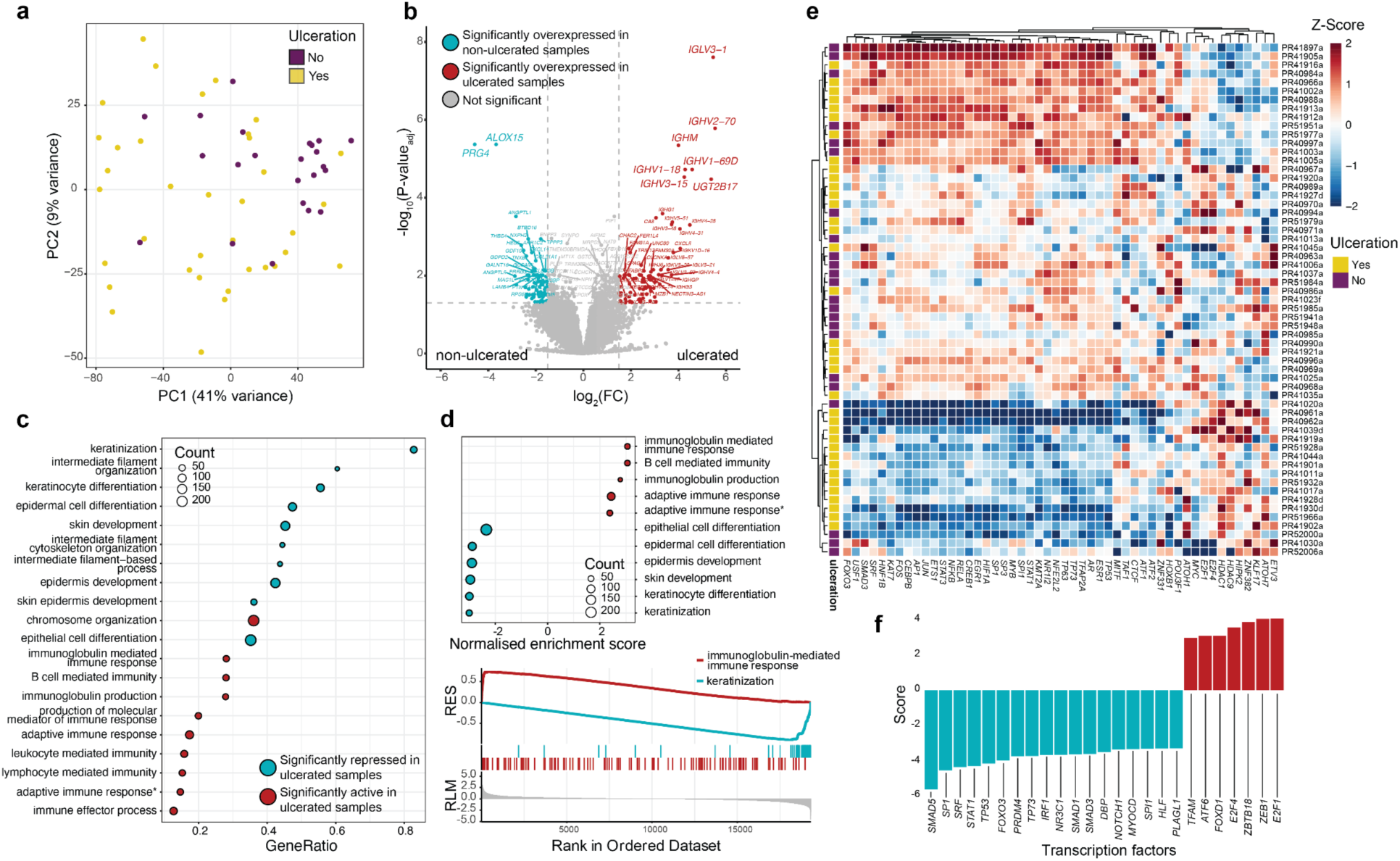
Transcriptional characterisation of acral melanoma tumours show an exacerbated humoral immune response and associated transcription factors. a) Principal component analysis of the 1000 most variable genes in all samples, stratifying by ulceration status, showing scores along the first (PC1) and second (PC2) principal components. b) Volcano plot showing differentially expressed between ulcerated and non-ulcerated samples. *P*-values are adjusted through the Benjamini-Hochberg method. FC: Fold change. c) Gene-set enrichment analysis (GSEA) displays the biological pathways more active in either ulcerated (red) and non-ulcerated (blue) samples by gene ratio. All processes depicted have *P*-values lower than or equal to 1.20x10^-10^. *Adaptive immune response based on somatic recombination of immune receptors built from immunoglobulin superfamily domains. d) Most differentially expressed terms between ulcerated and non-ulcerated samples by normalised enrichment score (top panel), GSEA plot showing the running enrichment score (RES) and ranked list metric (RLM) for the top enriched terms in ulcerated (red) and non-ulcerated (blue) samples (bottom panel). All processes in the top panel have *P*-values lower than 2.8x10^-16^. e) Heatmap showing the 50 most active transcription factors in the whole sample dataset, with an indication of sample ulceration status. f) Transcription factors that are significantly (corrected *P*-value < 0.05) active in ulcerated and non-ulcerated samples.

### RNA deconvolution analysis reveals plasma cells infiltration in ulcerated acral melanomas

Due to the immune cell involvement detected in gene expression and pathway enrichment analyses, we performed deconvolution of bulk RNA-seq data to estimate cell type abundance and evaluate the infiltration of immune cells in these AM samples (**Figure 3a**, **Supplementary Table 6**, **Methods**). We observed a statistically significant increase in plasma cells, M0 macrophages and eosinophils, along with increased expression of *CD79A*, the proinflammatory cytokine interleukin 6 (*IL-6)* and its receptor *IL6R* (**Figure 3b**), suggesting that inflammation and infection might accompany these lesions. No significant difference was found between other immune cells. Tissue immunostaining analyses on these samples identified increased protein expression of cell-surface major histocompatibility complex class II (MHCII) in ulcerated compared with non-ulcerated lesions (**Figure 3c**, **Methods**). These results indicate an ongoing antigen presentation process, which may be facilitated by B cells and macrophages. This shows the enrichment of immunoglobulin and B cell-mediated immunity in the ulcerated AM microenvironment, suggesting that an exacerbated humoral immune response and chronic inflammation may contribute to tumour progression^22,23^.

**Figure 3.**
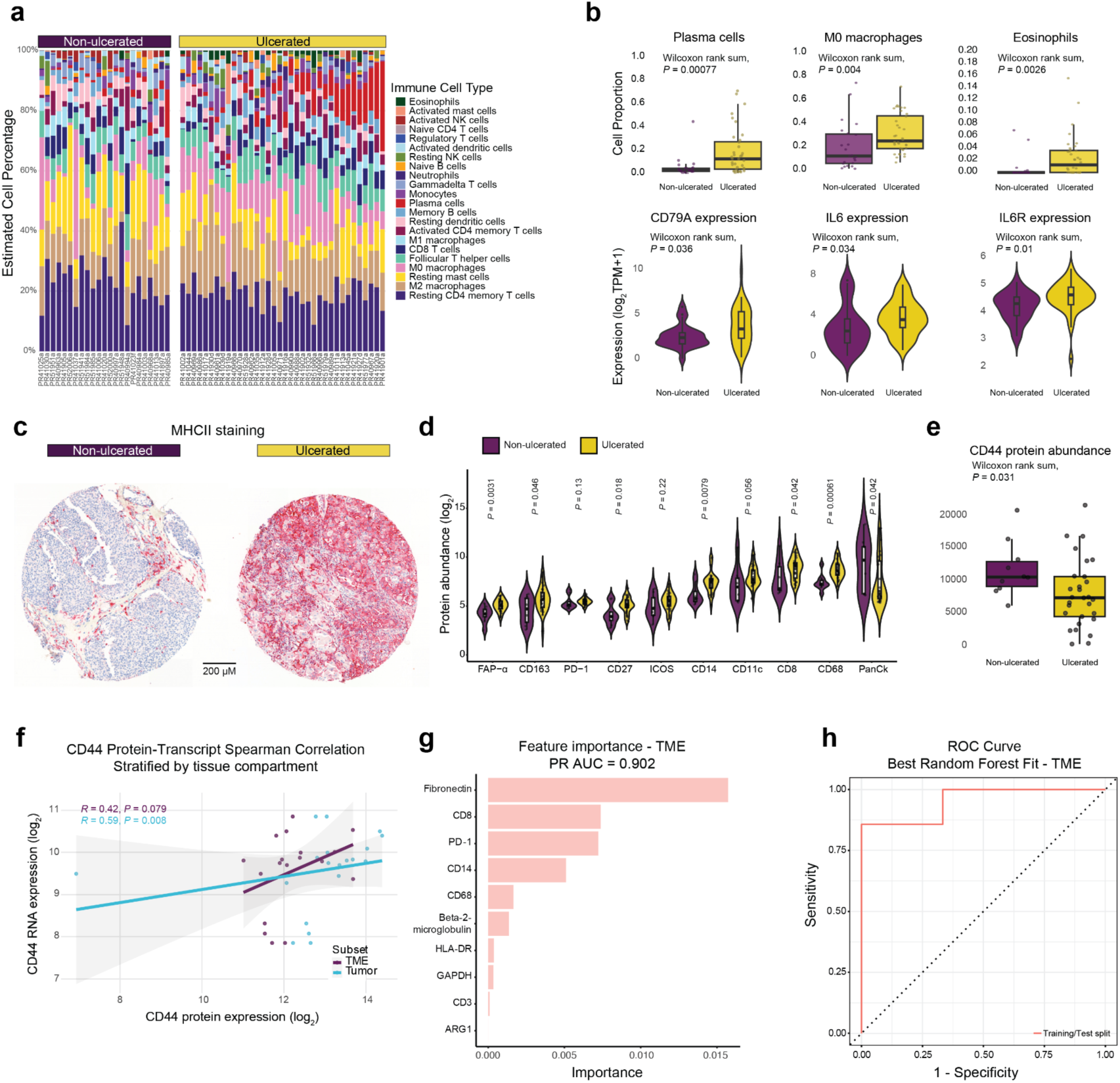
Plasma cells and inflammatory-response related genes are increased in ulcerated compared to non-ulcerated acral melanomas. a) Analysis of immune cell type composition in AM RNA-seq data using deconvolution (CIBERSORTx). Stacked bar plots illustrate the estimated proportions of immune cell types across the two clinical groups (ulcerated patients marked in yellow on the upper right, non-ulcerated in purple on the left). Red bars show plasma cell differences in the ulcerated group. b) Box plots depict cell types with significant differences between ulcerated and non ulcerated tumours: plasma cells, M0 macrophages and eosinophils. Violin plots display gene expression of *CD79A*, *IL6* and *IL6R* in ulcerated and non ulcerated tumours. c) Representative cores showing MHCII immunohistochemical staining (red chromogen) in ulcerated versus non-ulcerated lesions. Yellow indicates ulcerated patients, purple non-ulcerated patients. Scale bar, 200 μm. d) Violin plots comparing protein abundance differences in the TME, immune and stromal marker in ulcerated versus non ulcerated tumours. e) Box plot shows CD44 protein levels in ulcerated vs non ulcerated lesions. Wilcoxon rank-sum tests were used to evaluate significance. f) Spearman correlation between CD44 protein and transcript levels in the TME and tumour compartments. (Purple: TME, Blue: Tumour). g) Feature importance (permutation-based) utilising Random Forest Classifier. Bar plots show the most predictive proteins detected in the TME. h) Receiver operating characteristic (ROC) depicting the performance of the random forest in classifying ulceration within the TME model (PR AUC = 0.902, ROC = 0.762). The pink line shows the model’s sensitivity (true positive rate) plotted against 1-specificity (false positive rate) at various classification thresholds. The dotted diagonal line represents a random classification diagnostic value. N = 10 test cases. Yellow: Ulcerated, Purple: non ulcerated.

### Protein profiling and random forest analysis identifies robust myeloid presence and developing adaptive immune responses in ulcerated samples

To validate our findings from the transcriptome analyses and further characterise the presence of immune and stromal cell types in AM tissues, we profiled a tissue microarray containing a subset of these tumours and additional ones through spatial proteomic analysis using GeoMx DSP technology (**Methods**). This protein dataset comprised tumour cell (S100/Pmel17+) and tumour microenvironment/immune cell (TME, CD45+) regions from 45 distinct acral melanoma tumours (**Supplementary Tables 7-8**). Protein abundance analysis through unsupervised clustering identified a significant stromal signal in most samples. Fibronectin and alpha smooth muscle actin (SMA) were highly abundant at protein level (**Supplementary Figure 2**).

When comparing ulcerated versus non-ulcerated acral melanomas, we observed different protein marker profiles across the TME and tumour compartments. As expected, ulcerated lesions exhibited enhanced immune infiltration. Specifically, within the TME, ulcerated lesions displayed a significant increase in monocyte marker CD14, pan macrophage marker CD68 and macrophage M2 macrophage marker CD163, cytotoxic T cell marker CD8, lymphocyte co-stimulation marker CD27, and cancer-associated fibroblast marker FAP-α. We also observed upward trends in co-stimulatory receptor CD40 and dendritic cell marker CD11c (**Figure 3d**). Consistent with our transcriptomic analyses, the epithelial marker pancytokeratin exhibited significant downregulation in ulcerated samples (**Figure 3d**), indicating reduced epithelial characteristics in these lesions. Within the tumour compartment, we observed significant increase of myeloid cell markers (CD68, CD14, CD11c), alongside trends towards increased B cell infiltration (CD20, *P* = 0.069) and memory T cells-antigen experienced (CD45RO, *P*= 0.059) **(Supplementary Figure 2).** These findings collectively suggest enhanced myeloid cell presence and potential adaptive immune responses within ulcerated tumours. Additionally, the cell adhesion molecule CD44 was significantly decreased both at the RNA and protein level, suggesting altered cellular adhesion in ulceration (**Figures 3e,f**). Interestingly, we observed no significant differences in either tissue compartment for T cell activation marker ICOS or cytotoxicity marker granzyme B, indicating that while immune cell infiltration is increased in ulcerated tumours, the functional activation status of these cells requires further investigation.

We sought to identify proteins capable of distinguishing between ulcerated and non-ulcerated lesions, through the application of a random forest classifier (**Methods, Supplementary Table 9**). We attempted to determine the predictive proteins in both the tumour and TME compartments. Whereas the analysis in the tumour showed no predictive proteins, the TME model achieved strong predictive performance and well-balanced precision recall (area under the precision-recall curve = 0.92 and ROC-AUC = 0.762), indicating that the model significantly outperforms a random classifier (0.50). The top ranked proteins based on permutation feature importance were fibronectin, CD8, PD-1, CD14 and CD68 as the proteins that contributed most strongly to the model predictions (**Figures 3g,h**). These results were further supported by protein differential abundance analysis using a linear mixed model approach, which confirmed the significant overexpression of the monocytic/macrophage lineage within the TME compartment (CD68: *P*=0.025, FC=2.26; FAP-α: *P*=0.03, FC=1.87; CD14: *P*=0.046, FC=2.36, CD27, P = 0.046, FC = 2.01) (**Methods**, **Supplementary Figure 3**). Altogether, these findings indicate a robust myeloid presence and a predominance of an innate immune response, along with the emergence of adaptive immune responses that are likely predominantly humoral.

## Discussion

Ulceration is associated with a poor prognosis and is recognised as a risk factor for melanoma recurrence and metastasis^24,25^. Here, we sought to examine the molecular features of ulcerated AM tumours through a comprehensive analysis of transcriptome and proteome data, comparing ulcerated and non-ulcerated samples. These novel insights into the biological processes and unique immune microenvironment of ulcerated AMs not only enhance our understanding of this type of cancer, but may also inform therapeutic strategies targeting B cell-mediated immunity and inflammatory cascades in these aggressive tumours.

Our research found that ulcerated lesions exhibit a marked inflammatory profile and EMT-like features such as upregulation of the mesenchymal marker vimentin and downregulation of epithelial programs^26^. Some characteristics of the epithelial-to-mesenchymal (EMT) transition were observed; for example, EMT involves the loss of apical-basal polarity in epithelial cells, changes in the cytoskeleton, a decrease in cell-cell adhesion, and reorganization of cortical actin filaments in epithelial cells, among other features^27^ where EMT is linked to invasion and dissemination^28^. The transcription factor *ZEB1*, found to be more active in ulcerated samples (**Figure 2e**) is important for B cell differentiation and is considered a master EMT regulator, capable of inducing vimentin expression, which itself is a marker of EMT.

Immune cell infiltration is a key biomarker for clinical decision making in oncology^29,30^. We observed an inflammatory infiltrate in ulcerated lesions and confirmed an increased presence of monocytes/macrophages concordantly with previously reported results^12,31^. In various types of solid tumours, an increased concentration of macrophages is associated with a negative prognosis^32,33^. Given that our analyses also detected a significant increase in eosinophils, and considering the location of acral tumours (soles and heels), a response to infection is expected to play a role in these lesions. The upregulation of fibroblast activating protein (FAPα) and downregulation of CD44 at the protein level might reflect the tissue disruption and extracellular matrix (ECM) remodelling that is observed in ulcerated lesions. Immune-stromal interactions warrant further study due to its relevance in tumour development in chronic wounds^34^.

While most studies concentrate on the role of T lymphocytes, the functions of tumour-associated B cells (which present antigens to T cells via MHCII) and the humoral response are not well understood^35^. Poor prognosis in AM is thought to be related to a deficient immune response^36^ and ulcerated tumours have been reported to exhibit reduced tumour infiltrating lymphocytes (TILs), and a more proliferative phenotype^21^. In contrast, we observed upregulation of immunoglobulin-related mRNA transcripts that might be derived from B cells that differentiate into plasma cells upon antigen recognition. Notably, B cells act as professional antigen-presenting cells (APCs), being able to process and present BCR-bound antigens to CD4 T helper cells on MHC class II molecules^37^. At the protein level, we validated the expression of cell-surface major histocompatibility complex class II in ulcerated tumours, suggesting an elevated activity of antigen presentation that may result from the crosstalk between B and CD4 T helper cells. Macrophages might be involved as well. Additionally, CD20 and CD27, associated with B cell signaling and differentiation into plasma cells, were found to be increased in ulcerated tumours.

Plasma cells have been found in tumour infiltrates and can produce large amounts of cytokines and antibodies^38^. A research study discovered that class-switched B cells in the TME of melanoma tumours generated immunoglobulins, primarily IgG1, IgG2, and IgA, in comparison to healthy donors. The authors discovered evidence of autoantigen recognition and suggested an abnormal autoimmune-like reaction occurring more pronouncedly in patients with metastatic active disease^39^. In their study, Cabrita and collaborators^40^ discovered that B cells and plasma cells located in tertiary lymphoid structures (TLS) within melanoma tumours generated antibodies that target tumour-associated antigens. This finding was associated with improved responses to immunotherapy and underscored the potential significance of organized B cell responses in melanoma.

Although B cells can facilitate the elimination of tumour cells via phagocytosis, antibody-dependent cellular cytotoxicity (ADCC) and complement system activation, they can also contribute to the development and expansion of tumours^41^. Antibodies can be protumourigenic when they form circulating immune complexes with both tumour and non-tumour antigens, leading to chronic inflammation and tissue remodelling. This can ultimately result in immunosuppressive phenotypes^42^. Therefore, a better understanding of the profile of immunoglobulins produced in the TME, its functions and participating antigens would contribute to our understanding of tumour evolution and therapy response. Notably, the proinflammatory cytokine IL6, which transcript levels were upregulated in ulcerated lesions, is produced by tumours cells, antigen-presenting cells such as macrophages, dendritic cells, and B cells, as well as fibroblasts. IL6 can stimulate B cell differentiation and was initially discovered for its role in promoting antibody production^43^. The pleiotropic nature of IL6 in cancer has been further elucidated, with studies showing its important role in chronic inflammation, autoimmunity, and tumour progression^44–46^. Therapeutic approaches for IL6/IL6R blockade have been developed^47,48^.

The inflammatory profile of ulcerated tumours has been previously documented in cutaneous melanoma^21^. We hypothesise that the persistent tumour-promoting inflammation in ulceration, in addition to pathogen exposure in the microenvironment of acral ulcerated lesions, alters the local immune response, disrupting B cell immune tolerance mechanisms that lead to the emergence of autoimmunity features^39^. This then could result in feedback loops of exacerbated inflammation, tissue destruction (breakdown of the epidermal matrix) and impaired wound healing. On the other hand, tight junctions play a vital role in maintaining homeostasis, regulating the immune response, and promoting wound healing, with their barrier functions being of paramount importance. For instance, knockdown of claudin-1 induced a psoriasis-like condition in mice^49^. We hypothesize that loss of cell adhesion might follow the prolonged effect or tumour-promoting inflammation.

In the proteomic analysis, we could verify that although TME and tumour regions are efficiently separated in an unsupervised protein expression analysis, immune markers (e.g. CD14, CD68, CD20, CD11c) are detected in tumour regions. This fact probably reflects true immune infiltration in tumour regions (as confirmed through visual inspection of the immunofluorescence images). The prognostic importance of intratumoural immune cell presence within tumours has been documented^50,51^. For instance, CD14+ monocytes and tumour associated macrophages (TAMs) penetrate into tumours^52^ and complex mixtures of immune cells have been observed within melanoma nests. Additionally, tertiary lymphoid structures which include T and B cell conglomerates can develop within or adjacent to tumours^40,41^.

To the best of our knowledge, this is the largest study of the genomics and proteomics of ulcerated AM in a Latin American population. The limitations of this study include sampling bias since our Mexican cohort comprised more advanced tumours and insufficient numbers of thinner ulcerated lesions. AM is usually diagnosed at late stages in these populations^9,53^, and disparities in access to health care, poverty, lack of awareness, deficient access to trained dermatologists and oncologists, and an overburdened public healthcare system in Mexico exacerbate the problem of delayed presentation. More efforts are needed to ensure that clinical trial populations adequately reflect the diversity of patient populations around the world. We hope that efforts such as the one presented in this study help understand the molecular characteristics of ulcerated tumours that are detected at a later stage (which constitute the majority of the samples in this cohort), as well as contribute to diversifying the datasets available to the scientific community.

While our research revealed insights and correlations between molecular characteristics and ulceration, it is important to consider that these observations show a snapshot from a single moment in time. Consequently, the patterns we have identified may be both contributing factors to ulceration as well as reflect components of the wound healing response. Moreover, these patterns may overlap with elements of developing anti-tumour immunity and skin barrier defence (*i.e.* barrier breach and exposure to invading pathogens), therefore, establishing causality represents a challenge. The central question of whether these mechanisms precede or follow ulceration requires longitudinal and experimental studies.

Taken together, our results reveal an immunological landscape in ulcerated AM characterised by pronounced inflammatory signatures and a marked upregulation of the humoral immune response. Our analyses expand on previous investigations and contribute to a better understanding of the molecular features of ulceration in an understudied disease. We hope that studies such as this can help identify clinically useful biomarkers and eventually enable the detection of earlier lesions, consequently enhancing survival of AM patients worldwide, particularly in underserved populations where late-stage diagnosis remains prevalent.

## Methods

### Patient recruitment and sample collection

The protocol for sample collection was approved by the Mexican National Cancer Institute’s (Instituto Nacional de Cancerología, INCan, México) Ethics and Research committees (017/041/PBI;CEI/1209/17) and the United Kingdom’s National Health Services (NHS, UK) (18/EE/00076). Recruitment of patients and sample collection took place from 2017 to 2019. Patients attending follow up appointments at INCan that had previously been diagnosed with AM were offered to participate in this study, and upon signing a written consent form, were asked to provide access to a formalin-fixed paraffin-embedded (FFPE) sample of their tumour tissue that had been kept at the INCan tumour bank. This study included 59 patients for whom primary tumour material and clinical data was available. Patient IDs are not known to anyone outside the research group.

### RNA extraction and library preparation

RNA extraction from Formalin-Fixed Paraffin-Embedded (FFPE) tissue sections was performed at the Wellcome Sanger Institute (UK) using the AllPrep DNA/RNA FFPE kit. Steps included paraffin removal and RNA purification. The protocol yields purified total RNA that can be stored at -70°C for subsequent quality control analysis. Total RNA library preparation followed by exome capture using Agilent SureSelect AllExon v5 was performed. Transcriptome sequencing was performed on an Illumina HiSeq 4000 system with 150 bp paired end reads at the same institute.

### Transcriptome sequencing and processing

FastQC and multiQC were used for quality control of raw sequencing files. We performed exome-capture bulk RNA-seq on 146 total samples and after quality control, 86 samples remained for downstream analysis. From these, 59 samples were primary lesions that passed alignment quality control and had complete ulceration data, and were the ones reported in this study. Samples were discarded if their total read count fell below 25 million. Reads were mapped to the GRCh38 human reference genome using the splice aware aligner STAR, and counts were generated with HTSeq. Gene expression values were TPM normalized and subsequently underwent log2(TPM+1) transformation for downstream analyses.

### Differential gene expression analysis and GSEA

Differentially expressed genes (DEGs) were determined with DESeq2^54,55^ adjusting for Breslow depth, stage and batch. A gene was considered DE based on a Benjamini & Hochberg adjusted *P*-value < 0.05 and log2 Fold Change (LFC) ± 1.5. *P*-values were adjusted using Benjamini– Hochberg FDR correction for multiple testing^56^. GSEA was performed using clusterprofiler and the fgsea package^57^ with Gene Ontology (BP, MF, CC), molecular signature database (MSigDB) and KEGG pathways.

### Transcription Factor Activity Inference

TF activity inference was performed using decoupleR^58^ which is based on a univariate linear modelling (ULM) and prior knowledge networks (GRNs). We used the CollecTRI Database^59^ that includes a curated assemblage of transcription factors (TFs) and their corresponding transcriptional targets (sourced from 12 distinct repositories). The interactions within CollecTRI are assigned weights based on their regulatory mechanism (activation or inhibition). Transcription factor activities were obtained from the t-values of the differentially expressed genes between ulcerated and non ulcerated tumours.

### RNA deconvolution analysis

Immune cell composition analysis of acral melanoma samples was performed using the CIBERSORTx digital cytometry algorithm^60^. The analysis was run in absolute mode to estimate cell fractions with 100 permutations. Expression data were normalized to transcripts per million (TPM) before deconvolution. Results were visualized and compared between ulcerated and non-ulcerated samples to identify significant differences in immune cell composition.

### Tissue microarray assembly and histology

Acral FFPE samples were used to build a 152-core tissue microarray (TMA), comprising 150 tumour cores from acral melanoma patients plus two control tissue cores. Each patient contributed between 1-4 cores depending on tissue availability and integrity. This TMA underwent immunohistochemical staining with H&E and MHCII (LGII.612-14 antibody). The Leica Bond Polymer Refine Red Detection kit (7730-DS9390) was used. Slides were digitized at the University Hospital Zurich using an Anperio Scanscope microscope with a 20x objective. QuPath^61^ was used for visual inspection and quantification of positively stained cells.

### Proteomic analysis

Serial sections from the TMA were used for GeoMx DSP profiling. H&E-stained slides were scanned at 20x and used to guide ROI selection. Representative histological regions of interests (ROI) from the TMA cores were selected using morphology markers to identify CD45 (leucocytes), S100B/Pmel17 (tumour cells) and SYTO13 (nuclei), by a dermatopathologist in conjunction with members of the research team. ROIs were selected based on tissue integrity, architecture, and balance of clinical characteristics. Antibody panels were used to identify cell lineage of immune cells, stromal cells, endothelial cells and epithelial cells. The GeoMx DSP protein assay was performed according to the manufacturer’s instructions (Nanostring, Seattle, WA). The data underwent quality control and was normalised based on the negative controls and subsequently log2-transformed. Z-scores were used for clustering and visualisation.

### Random forest analysis

Four machine learning models were tested using the R Tidymodels package: random forest, nearest neighbours (knn), logistic regression and support vector machine (svr). Random Forest showed the best predictive performance and was therefore selected. Separate models for each tissue compartment (tumour and TME) were trained using stratified five-fold cross-validation and grid search-based hyperparameter tuning. The Area under the Precision-Recall curve (PR AUC) and ROC were used for model evaluation. Permutation-based importance scores were generated to evaluate feature’s contribution to model performance.

### Linear Mixed Effects

To determine differentially expressed proteins in ulcerated vs non ulcerated tumours accounting for repeated measurements (multiple ROIs per patient), a linear mixed effects model was applied for each protein, setting ulceration status as fixed effect and patient as random effect. Log2 transformed protein counts were used. The effect size was estimated (Log2 fold changes) and the Benjamini-Hochberg procedure was applied to control false discovery rate (FDR < 0.1).

### Statistical analyses

A *P*-value of < 0.05 was considered statistically significant. *P-*values were derived from a two-tailed Wilcoxon rank sum test. In both the text and figures, we have included false discovery rate (FDR) adjusted p-values. All analyses were performed using R (version 4.3.2) and Bioconductor version 3.18 (BiocManager 1.30.22). Visualizations were made with ggplot2.

## Code Availability

Sequencing data are available at the European Genome-Phenome Archive (EGA) under ENA accession number EGAS00001003758. Protein data and code is available at https://github.com/CGBio-Lab/Ulceration_Acral_Melanoma.

## Acknowledgements

We express our sincere gratitude to all the patients and their families for their participation and for their invaluable contribution towards enhancing understanding of acral melanoma in Mexico. We are also thankful to members of the CGBio lab team at LIIGH-UNAM for valuable discussions regarding the findings in this article. The authors wish to thank Luis A. Aguilar, Alejandro de León and Carlos S. Flores from the Laboratorio Nacional de Visualización Científica Avanzada and Jair S. García Sotelo, Abigayl Hernández, Eglee Lomelín, Iliana Martínez, Rebeca Muciño, María A. Ávila, Alejandra Castillo, Carina Uribe Díaz and Christian Molina-Aguilar from the International Laboratory for Human Genome Research, National Autonomous University of Mexico. Work included in this paper has been funded by Wellcome Trust (204562/Z/16/Z and 227228/Z/23/Z to C.D.R.-E.), the Melanoma Research Alliance (Pilot Award #825924, to C.D.R.-E.), the Mexican National Council of Humanities, Science and Technology (CONAHCYT, FOSISS A3-S-31603, to C.D.R.-E.), Programa de Apoyo a Proyectos de Investigación e Innovación Tecnológica (PAPIIT UNAM) (IN209422 to C.D.R.-E.), and the Wellcome Sanger Institute through an International Fellowship. This project was also supported by the MRC Dermatlas project; MR/V000292/1. M.E.V.-C. is a doctoral student from the Programa de Doctorado en Ciencias Biomédicas, Universidad Autónoma de México (UNAM) and has received a CONAHCYT fellowship with number 521002192. The funders had no role in the study design, data analysis, or preparation of the manuscript.

## Ethics statement

The protocol for sample collection was approved by the Mexican National Cancer Institute’s (Instituto Nacional de Cancerología, INCan, México) Ethics and Research committees (017/041/PBI;CEI/1209/17) and the United Kingdom’s National Health Services (NHS, UK) (18/EE/00076).. All patients signed informed consents to participate in this study.

## Competing interests

JSR reports in the last 3 years funding from GSK and Pfizer & fees/honoraria from Travere Therapeutics, Stadapharm, Astex, Owkin, Pfizer, Grunenthal, Tempus and Moderna. The authors declare no competing interests.

## Supplementary Figures

**Supplementary Figure 1.**
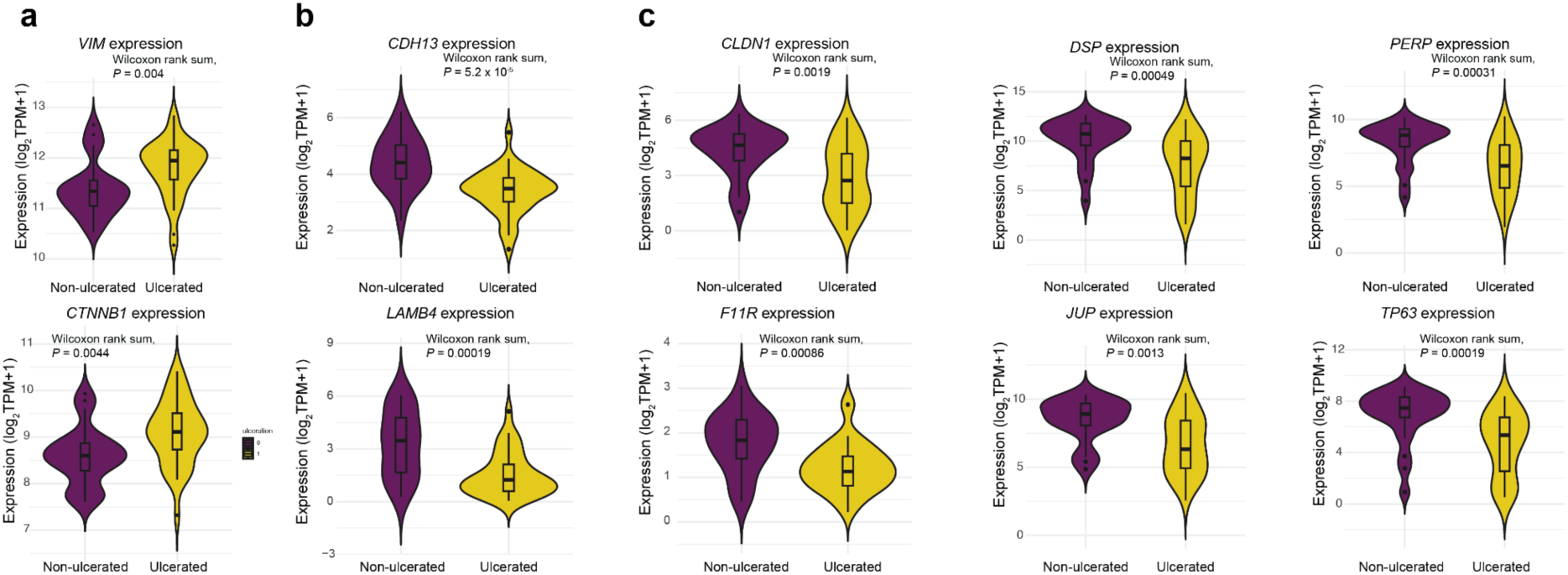
Violin plots show the expression levels of cell adhesion genes in ulcerated and non-ulcerated samples. a) Genes related to epithelial-to-mesenchymal transition. b) Genes related to intercellular junctions. c) Genes related to desmosomal components.

**Supplementary Figure 2.**
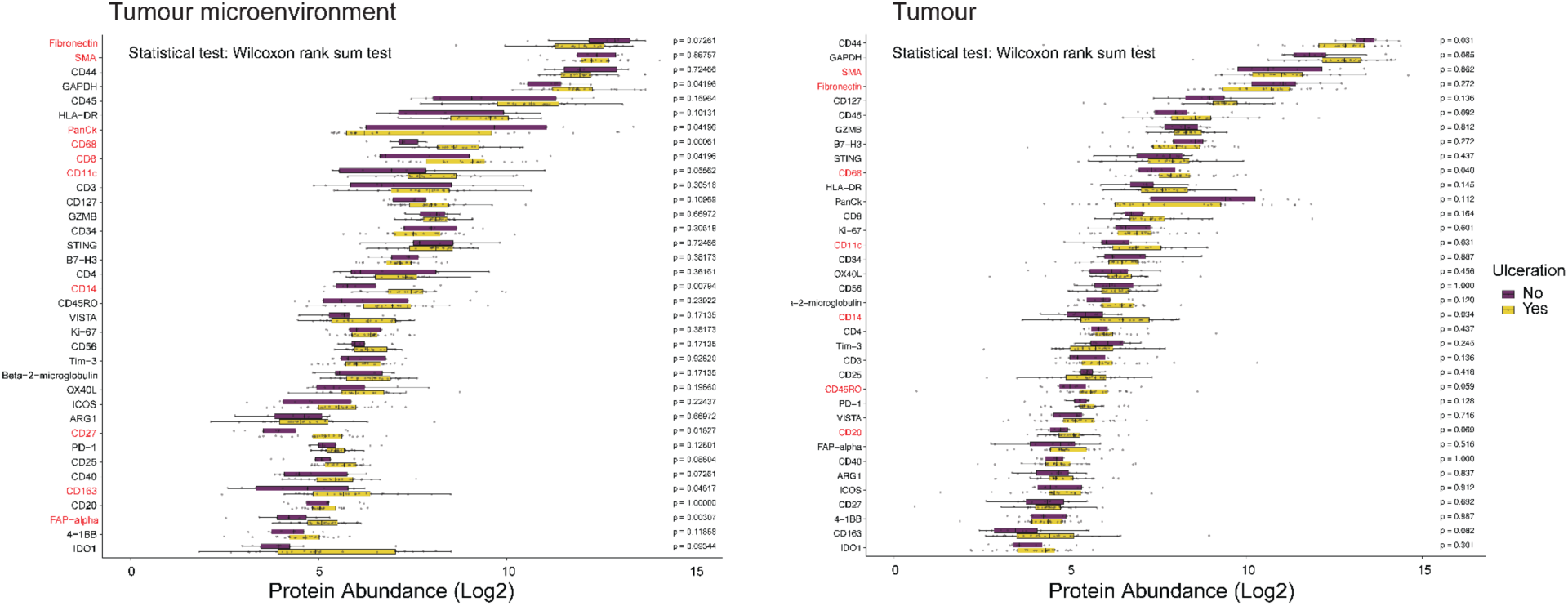
Comparison of the expression levels of diverse proteins in ulcerated and non-ulcerated samples. a) Quantification in the tumour microenvironment compartment. b) Quantification in the tumour compartment.

**Supplementary Figure 3.**
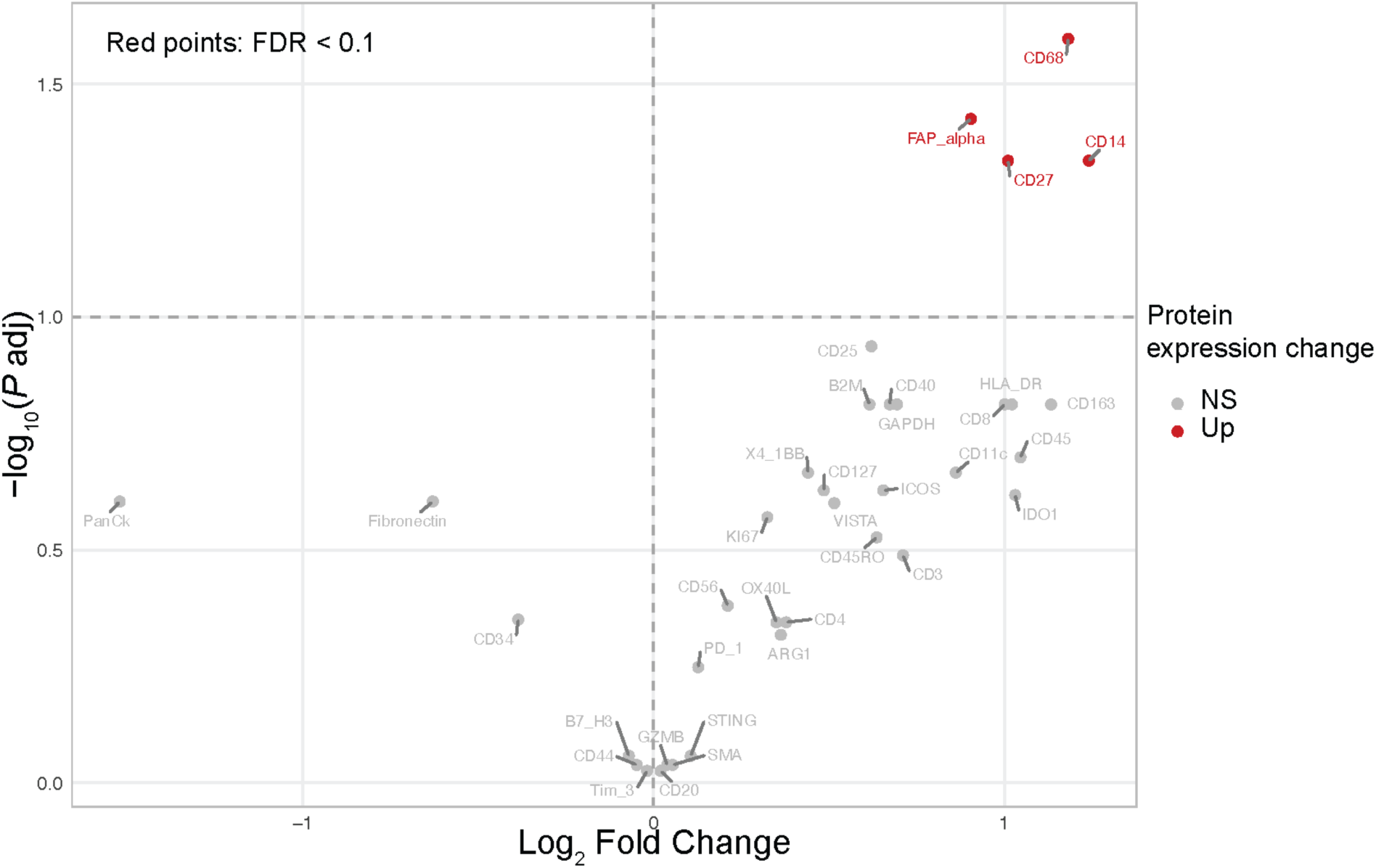
Volcano plot showing differences in protein levels within the tumour microenvironment compartment between ulcerated and non-ulcerated samples, analyzed using linear mixed models (LMMs). Proteins that are significantly overexpressed in ulcerated lesions are represented by red dots.

## Notes

### Author Declarations

The protocol for sample collection was approved by the Mexican National Cancer Institute's (Instituto Nacional de Cancerologia, INCan, Mexico) Ethics and Research committees (017/041/PBI;CEI/1209/17) and the United Kingdom's National Health Service (NHS, UK) (18/EE/00076). All patients signed informed consent.

