## Supplementary figures and images for "The microenvironment of ulcerated acral melanoma is characterised by an inflammatory milieu and an enhanced humoral immune response"

### Supplementary Figure 1

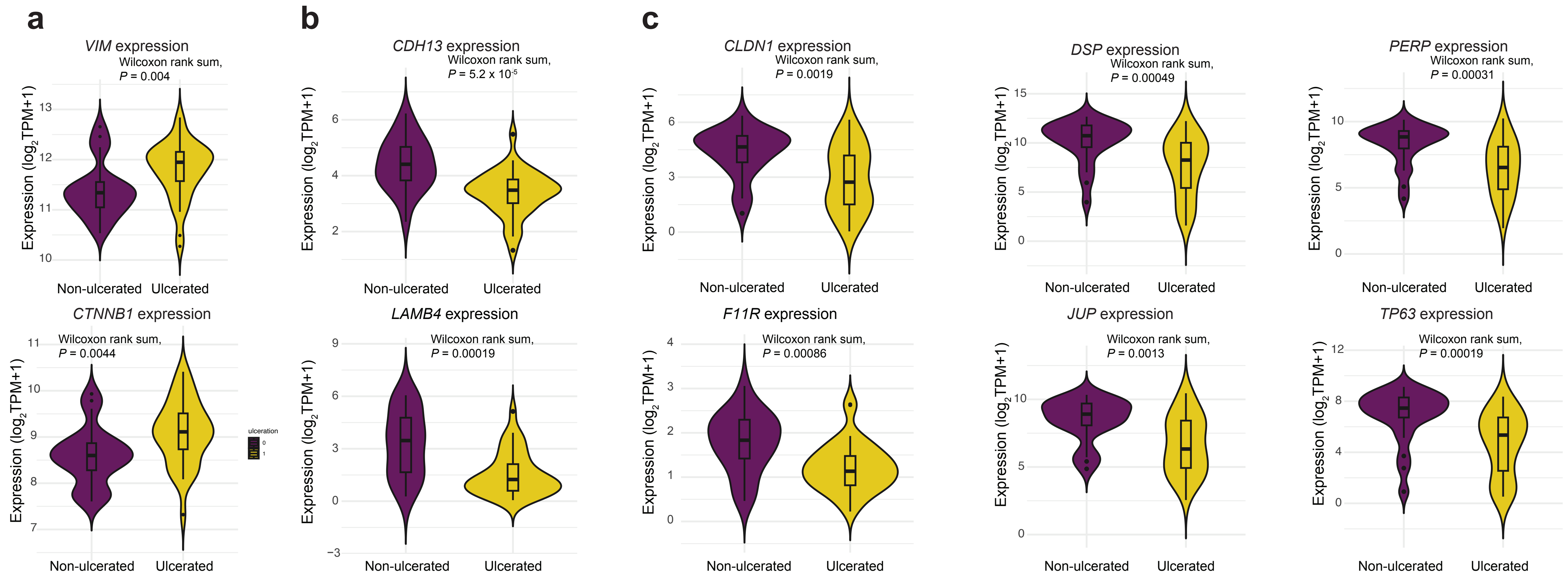

### Supplementary Figure 2

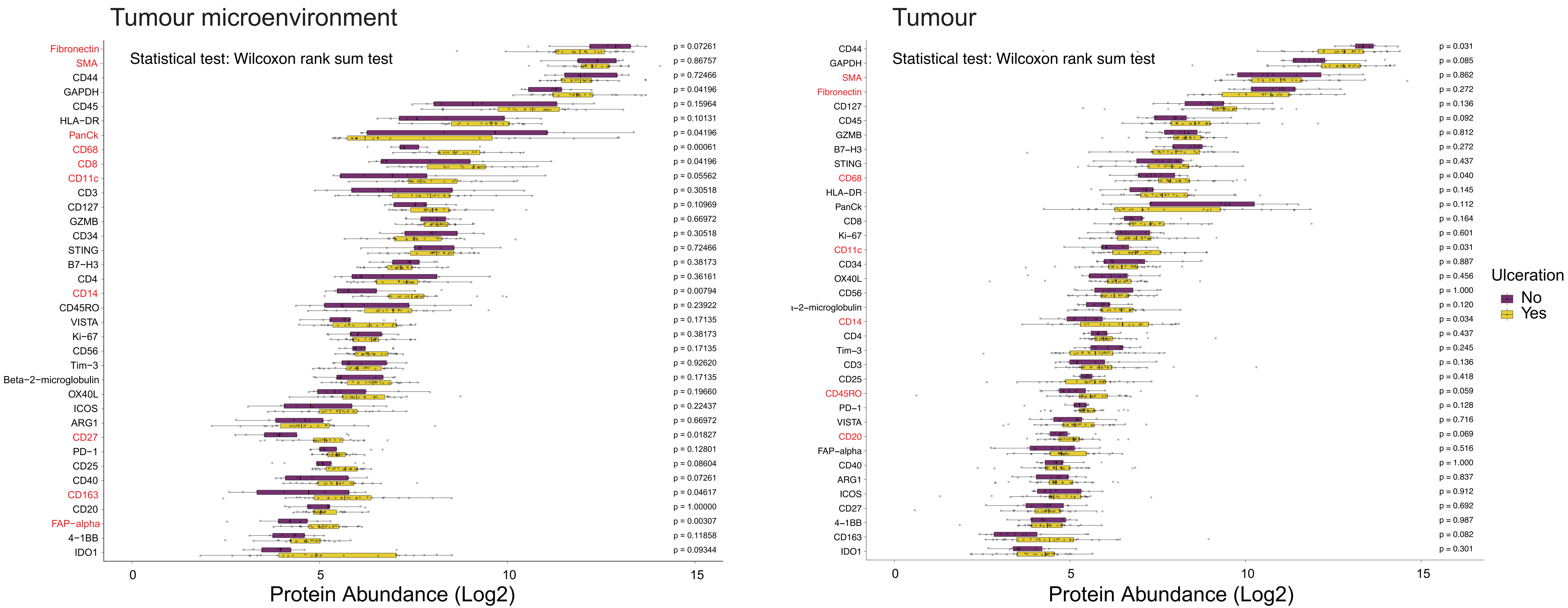

### Supplementary Figure 3

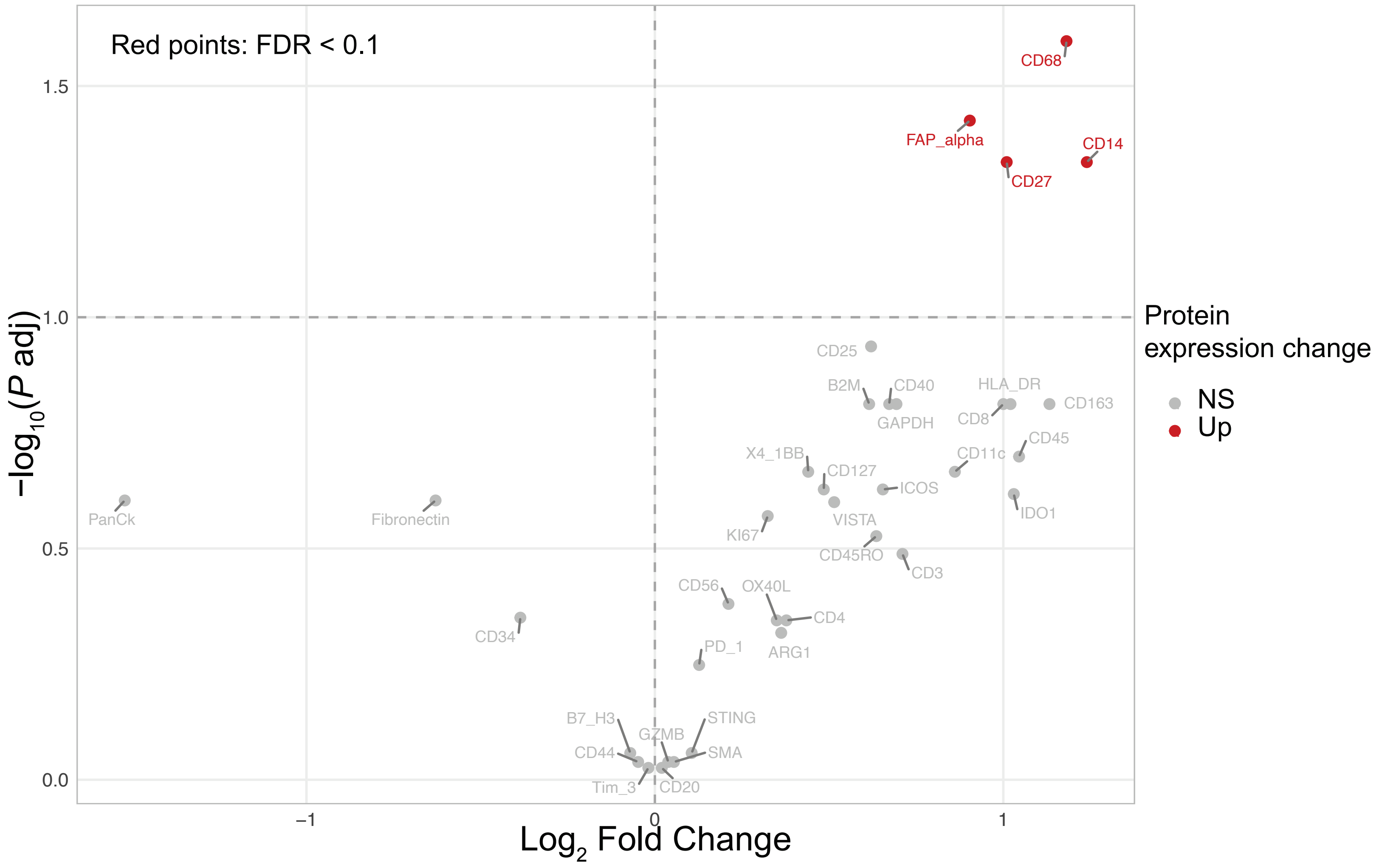
